# Tasks for assessing dystonia in young people with cerebral palsy

**DOI:** 10.1101/2025.09.07.25334597

**Authors:** Emma Lott, Alyssa Rust, Leon Dure, Darcy Fehlings, Toni Pearson, Roser Pons, Jonathan W. Mink, Bhooma R. Aravamuthan

**Author notes:** Address correspondence to: Bhooma R. Aravamuthan, MD, DPhil, Division of Pediatric Neurology, Department of Neurology, Washington University School of Medicine, 660 South Euclid Avenue, Campus Box 8111, St. Louis MO 63110-1093.

## Abstract

**Background:** Dystonia in childhood is typically associated with cerebral palsy (CP). Dystonia severity scales for CP require prolonged exam protocols with numerous tasks, often making them onerous for routine clinical use.

**Objective:** To identify which individual tasks best approximate dystonia severity compared to the gold standard full protocol.

**Methods:** In this cross-sectional study, comprehensive exam protocol videos were taken during routine care of ambulatory people with CP age 5 and up. Five pediatric dystonia experts reviewed individual tasks and the full protocol for dystonia using the Global Dystonia Severity Rating Scale. Experts’ written scoring justifications were qualitatively analyzed to determine commonly cited features of dystonia.

**Results:** When examining the difference in dystonia severity ratings between each task and the full protocol, seated upper extremity tasks had the lowest variance (p<0.05, F-test) and had lower score differences than the stand/walk/run and seated lower extremity tasks (p<0.05, repeated measures Friedman test). Experts most commonly identified the following movements as dystonic: wrist flexion (8.4% of all movement statements), finger flexion (7.3%), wrist ulnar deviation (6.8%), toe dorsiflexion (8.4%), ankle inversion (7.9%), and ankle plantarflexion (6.4%). Experts rated dystonic movements as more severe if they were consistently triggered by multiple stimuli (26.8% of all severity statements) or functionally impactful (20.7%).

**Conclusions:** Seated upper extremity tasks may be valuable for identifying dystonia and estimating its severity during routine clinical care. Clinical dystonia severity assessment could be guided by assessing specific dystonic movements, the consistency with which they are triggered, and their functional impact.

---

Cerebral palsy is the most common motor disability across the lifespan and is commonly associated with dystonia, a debilitating, frequently painful, and often treatment refractory movement disorder.^1–3^ Dystonia in CP affects up to 2 of every 1000 people in the US, comparable to the prevalence of Parkinson’s disease.^3–5^ However, despite its high prevalence and high impact, there are no data-driven treatment paradigms for dystonia in CP.^6,7^ Developing data-driven treatments requires the ability to assess dystonia severity in a standardized way across centers to facilitate large scale clinical trials. However, current dystonia severity assessments have been criticized for requiring prolonged video recordings or in person assessments (often lasting more than 30 minutes).^8,9^ This can be both expensive (requiring many personnel hours) and exhausting for people with CP to execute. Furthermore, the longer these severity assessments take, the less feasible they are to execute during routine clinical care, making it difficult to clinically assess the impact of dystonia treatments. Therefore, of the many tasks currently used across dystonia severity assessments, it would be valuable to determine the subset of tasks most useful for assessing dystonia in young people with CP.

Gold standard dystonia assessment is expert consensus review of neurologic exam videos.^10–12^ We have shown that experts most commonly refer to the seated alternating hand open-close task when reviewing videos for arm dystonia, particularly when attempting to differentiate between the absence of dystonia and the presence of mild dystonia.^12^ We have also shown that experts cite leg adduction variability and amplitude when assessing gait videos for leg dystonia and that these features quantifiably track with expert assessed leg dystonia severity.^10,11^ Therefore, across all tasks included in current dystonia severity assessments, a subset of tasks (like seated upper extremity tasks and walking) may be particularly valuable for dystonia assessment.

Our objective was to explore what might be a valuable subset of tasks that can be used to identify dystonia and assess its severity during routine clinical care of a young person with CP. We asked international experts in dystonia assessment (LD, DF, TP, RP, JWM) to review videos of young people with CP for dystonia and discuss these videos to reach consensus agreements about dystonia severity. We had them review individual tasks for each subject with CP and then finally review the full protocol, separated by 1-2 months between video reviews to limit recall bias. We then compared their dystonia severity ratings for each individual task with their severity ratings when reviewing the full protocol (which we considered to be gold standard assessment) to see which task grouping came closest to the dystonia ratings from the full protocol. We also assessed what movement and severity features experts cited when assessing dystonia. Using this methodology, our goals were to: 1) Identify the shortest subset of tasks that could be used to identify dystonia and assess dystonia severity comparably to expert review of the full protocol, and 2) Describe the features dystonia experts look for when identifying dystonia and assessing its severity.

## Methods

We received Washington University School of Medicine Institutional Review Board approval for this project (Approval Number: 202306016; Date of Approval 06/05/2023).

### Subjects

Subjects were recruited from the St. Louis Children’s Hospital Cerebral Palsy Center between 12/1/2022 and 1/31/2023 if they were diagnosed by a Center clinician as having CP per the 2006 definition,^13^ were independently ambulatory (so that they could complete all tasks including standing, walking, and running), had spasticity documented as their predominant tone pattern (to force experts to identify features that distinguished dystonia from co-existing spasticity), and if they were at least 5 years old (to increase the developmental likelihood that the person’s attention span would allow for the completion of a prolonged video protocol). Spasticity and dystonia were identified in each subject by the treating clinician during the clinic visit using the Hypertonia Assessment Tool.^14^ Of note, the presence or absence of dystonia was not used as a part of the inclusion criteria noting that the prevalence of dystonia in independently ambulatory people with CP and spasticity is between 50-60%.^10,11,15^ Subjects were excluded if they could not attempt completion of all video tasks (i.e. if they were behaviorally, attentionally, or cognitively unable to attempt the tasks as determined by the caregiver or the subject). Videos were obtained as a part of clinical care.

### Video protocol

Tasks were aggregated from three dystonia severity assessments validated in people with CP: the Barry Albright Dystonia Scale (BADS),^16^ the Dyskinetic Cerebral Palsy Impairment Scale (DIS),^17^ and the Movement Disorders – Childhood Rating Scale (MD-CRS).^18^ To protect participant identity during expert review, the heads of the participants were blurred (ShotCut, Meltytech, LLC) and face/speech tasks were not included. Tasks were divided into four groups:

1. Lying down tasks
  a. Reaching for an object
  b. Rolling to one side
2. Seated upper extremity tasks
  a. Alternating hand open-close
  b. Rapid finger tapping
  c. Reaching for an object
3. Seated lower extremity tasks
  a. Moving foot in a circular motion
  b. Foot tapping (unsupported, without a stool)
  c. Foot tapping (supported, with a stool)
4. Standing, walking, and running

In addition to the tasks outlined above, each task grouping also included a period of relative rest where subjects were asked to not move. Of note, the only true period of rest was during the lying down task where the body was fully supported. Relative rest during the seated and upright tasks required some degree of active support of the body. The full video protocol is available in the Supplementary Methods. Videos were recorded on tripod-mounted Google Pixel 3a smartphones at 1920×1080 pixel resolution at 30 frames/second. For lying down videos, the person was lying on a height adjustable bed with the height adjusted to be as close to the floor as possible while the smartphone was taped to the ceiling. This allowed for the full body to be in frame during the recording.

### Expert consensus building and rating

Experts separately reviewed each of the four task groupings plus the full protocol (five individual review periods in total) from 6/1/23 to 3/15/24 for all recruited subjects. Review periods were separated by 2 months to reduce the likelihood that experts would recall their previous scores on one task grouping and use that to inform their subsequent scores on other task groupings. For each of these five review periods, experts first independently reviewed videos for dystonia severity in the arm, trunk, and leg using the Global Dystonia Rating Scale (GDRS), which does not prescribe specific tasks or metrics for assessing dystonia severity and which we have previously established has high inter-rater reliability for dystonia assessment in people with CP.^11,19^ Experts were asked to provide free text explanations to justify what aspects of the video prompted them to say that there was or was not dystonia and what movement qualities helped them gauge dystonia severity using the GDRS. Expert assessments were entered in a REDCap survey. If there were subjects where the total GDRS score discrepancy between experts was greater than 10 points, their videos were reviewed by all experts in virtual consensus building meetings over Zoom. Of note, the GDRS is a 10-point Likert grading scale (0-no dystonia, 5-moderate dystonia, 10-severe dystonia) in each body segment assessed. For these video reviews, noting that face and neck dystonia were not rated, the total possible GDRS score was 90 (10 points each for the bilateral proximal and distal arms and legs and an additional 10 points for the trunk). A median whole body GDRS of 1 or more across all 5 experts indicates that at least 3 out of 5 experts agreed that the person in the video displayed dystonia. In the virtual consensus building meetings, experts with discrepant scores were asked to talk through how they scored the video and all experts were then invited to comment. After this discussion, experts were asked to re-score the video and submit these revised scores via a REDCap survey. When available, the revised scores were used for further analysis. Experts’ GDRS scores were separately tabulated for each subject for each task grouping for the full body (possible score range 0-90), arm subscore (possible range 0-40), and leg subscore (possible range 0-40). Assessments of the arm and leg subscores were used to determine whether there was a regional difference in dystonia assessment between tasks and to ensure adequate assessment of people with focal or segmental dystonia. The differences between GDRS scores for each task grouping and the corresponding scores for the full protocol were calculated.

### Qualitative analysis

Experts’ free text descriptions were analyzed using a conventional qualitative content analysis approach to break down the content into salient codes.^20^ The transcripts were coded by two independent coders (EL, AR) blinded to the severity scores of the participants. Both coders met to resolve any discrepancies by consensus discussion and consolidated their codes into a single code book. Codes that were cited at least twice were included in further analysis and divided into four categories: codes describing dystonic movement features in the upper extremities, lower extremities, and trunk, and codes describing how experts approached grading dystonia severity.

### Statistical analysis

All statistical analyses were done in GraphPad Prism (version 10, GraphPad Software LLC). F-tests with Bonferroni corrections were used to determine the homogeneity of variances between GDRS score differences across task groupings (lying down, seated upper extremity, seated lower extremity, and stand/walk/run). If variances were non-homogeneous across groups, a repeated measures Friedman test and post-hoc Dunn’s test for multiple comparisons were used to compare score differences between the task groupings. If variances were homogeneous, a repeated measures ANOVA with Geisser-Greenhouse correction and post-hoc Dunnet’s test for multiple comparisons were used.

Within each code category (upper extremity, lower extremity, trunk, and severity), expert-cited code frequencies were compared between task groupings using a Chi-square test with Yates correction followed by post-hoc Chi-square tests with Bonferroni correction for multiple comparisons.

Significance level was set a priori at p<0.05.

### Data Availability

Anonymized data will be made available by request from any qualified investigator.

## Results

Out of the 112 subjects that met inclusion criteria, 10 subjects agreed to record the full video protocol as a part of their clinical care and 7 were able to complete recording the full video protocol. For these 7 subjects, the full video recording process (including set up and transit between the clinic room and hallway for different parts of the video recording) took 45 minutes to 1 hour. The video recordings in isolation were 24-28 minutes long in total.

### Comparison of GDRS scores between individual task groupings and the full protocol

When directly comparing GDRS scores between each task grouping and the full protocol, neither the lying down task grouping nor the seated upper extremity task grouping yielded total body scores, arm scores, or leg scores that were significantly different from the full protocol (Table 1, Figure 1A, 1C, 1E). The seated lower extremity task grouping had significantly higher total body GDRS scores (p=0.03, Friedman test) and leg GDRS scores (p=0.005, Friedman test) compared to the full protocol. The stand/walk/run task had significantly higher total body GDRS scores compared to the full protocol (p=0.04, Friedman test) (Figure 1A, Table 1). That is, raters identified more severe dystonia when rating the lower extremity tasks or stand/walk/run tasks in isolation than when rating all tasks together in the full protocol.

**Table 1.** Summed Global Dystonia Severity Rating Scale (GDRS) scores for the body (arms+legs+trunk), arms, and legs compared between task groupings.

|  | GDRS score for each subject (median, range) |  |  |  |  |  |  | Adjusted<br>*p* |
| --- | --- | --- | --- | --- | --- | --- | --- | --- |
|  | 1 | 2 | 3 | 4 | 5 | 6 | 7 |  |
| **Total Body** |  |  |  |  |  |  |  |  |
| Lying down | 0, 0-4 | 0, 0-0 | 15, 12-18 | 4, 0-8 | 2, 0-15 | 0, 0-8 | 13, 10-16 | 0.8 |
| Sitting – UE | 2, 0-11 | 0, 0-3 | 3, 0-9 | 1, 0-6 | 3, 0-4 | 0, 0-8 | 14, 5-17 | 1 |
| Sitting – LE | 8, 0-8 | 1, 0-8 | 8, 6-8 | 2, 0-5 | 15, 13-19 | 4, 0-9 | 17, 12-27 | 0.03* |
| Stand/Walk/Run | 4, 3-6 | 2, 0-6 | 18, 11-23 | 6, 2-10 | 10, 9-16 | 0, 0-0 | 0, 0-14 | 0.04* |
| Full Protocol | 1, 0-7 | 0, 0-5 | 7, 4-9 | 0, 0-6 | 2, 0-10 | 0, 0-2 | 10, 10-14 | - |
| **Arm** |  |  |  |  |  |  |  |  |
| Lying down | 0, 0-0 | 0, 0-0 | 4, 3-8 | 1, 0-4 | 2, 0-7 | 0, 0-2 | 8, 7-11 | 1 |
| Sitting – UE | 0, 0-3 | 0, 0-0 | 2, 0-4 | 0, 0-2 | 0, 0-3 | 0, 0-0 | 6, 5-8 | 1 |
| Sitting – LE | 4, 0-6 | 0, 0-0 | 0, 0-3 | 0, 0-1 | 6.5, 5-9 | 0, 0-1 | 5, 4-7 | 1 |
| Stand/Walk/Run | 4, 3-6 | 1, 0-2 | 7, 3-12 | 3, 2-8 | 5, 3-6 | 0, 0-0 | 0, 0-4 | 0.08 |
| Full Protocol | 1, 0-2 | 0, 0-1 | 5, 4-7 | 0, 0-2 | 0, 0-4 | 0, 0-0 | 6, 4-8 | - |
| **Leg** |  |  |  |  |  |  |  |  |
| Lying down | 0, 0-4 | 0, 0-0 | 9, 6-15 | 3, 0-5 | 0, 0-7 | 0, 0-6 | 4, 2-6 | 0.6 |
| Sitting – UE | 2, 0-8 | 0, 0-3 | 3, 0-5 | 0, 0-4 | 0, 0-3 | 0, 0-8 | 8, 0-9 | 1 |
| Sitting – LE | 2, 0-8 | 1, 0-8 | 6, 5-8 | 2, 0-4 | 8.5, 8-9 | 2, 0-9 | 12, 8-18 | 0.005* |
| Stand/Walk/Run | 0, 0-0 | 1, 0-4 | 7, 4-14 | 1, 0-4 | 6, 3-12 | 0, 0-0 | 0, 0-8 | 1 |
| Full Protocol | 0, 0-6 | 0, 0-4 | 0, 0-4 | 0, 0-4 | 2, 0-5 | 0, 0-2 | 5, 2-8 | - |

\* $p < 0.05$ for repeated measures Friedman test and post-hoc Dunn's test for multiple comparisons
between each task grouping and the full protocol. UE – upper extremity. LE – lower extremity.

**Figure 1.**
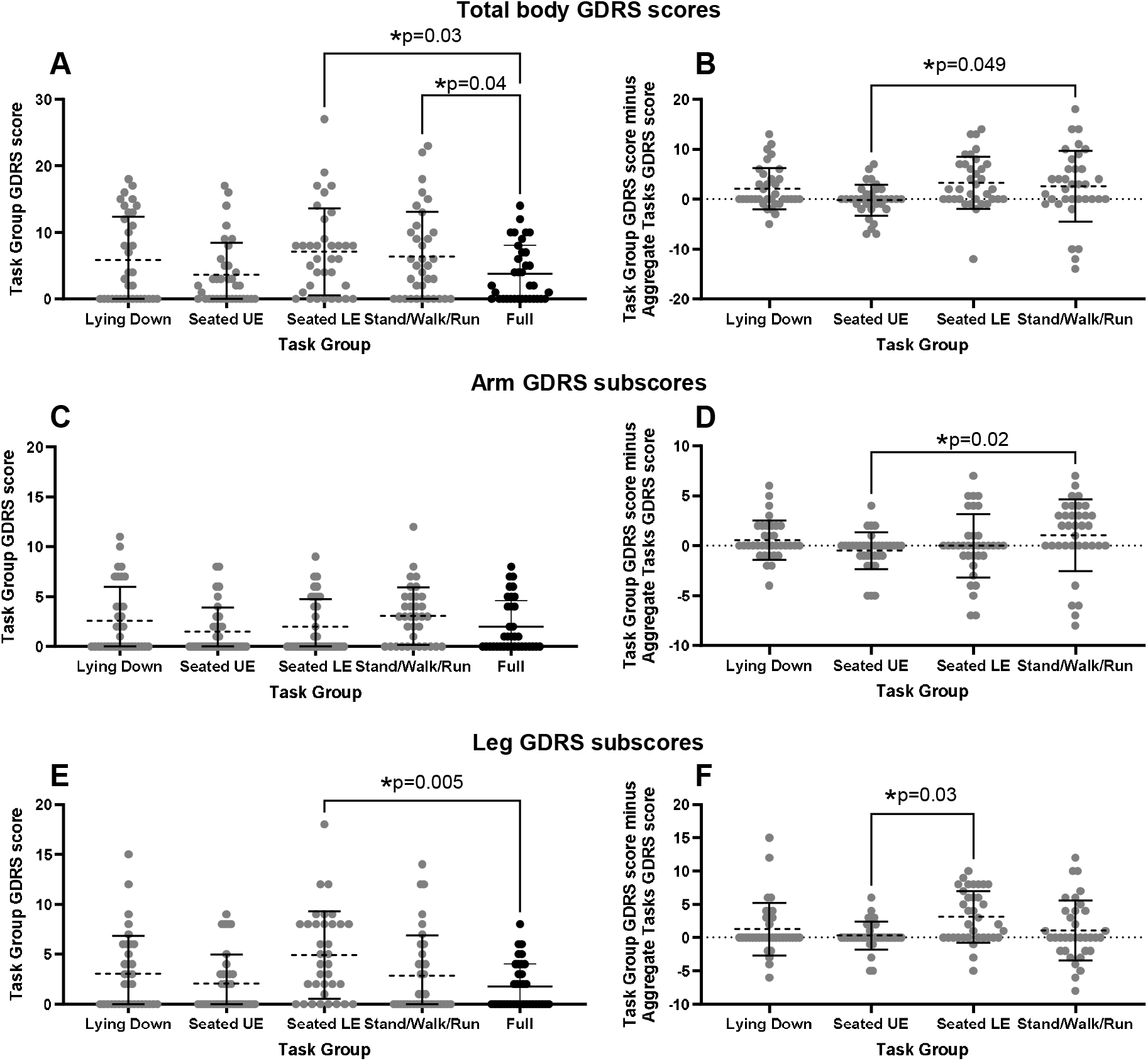
Comparison of Global Dystonia Severity Rating Scale (GDRS) scores across tasks. Data points are GDRS scores or score differences per expert rater per subject. *p<0.05 for repeated measures Friedman test and post-hoc Dunn’s test comparing each task grouping and the full protocol (left) or comparing task groupings (right). U/LE – upper/lower extremity.

We next subtracted the GDRS scores for each task grouping from the corresponding GDRS scores for the full protocol (zero values indicate that the task grouping score matched the full protocol score) (Table 2, Figure 1B, 1D, 1E). We compared these score differences between task groupings. The seated upper extremity task grouping yielded total body, leg, and arm GDRS scores that most frequently matched the corresponding full protocol GDRS score (5-6 out of 7 subjects with GDRS scores matching the corresponding full protocol score) compared to all other task groupings (2-5 out of 7 subjects with GDRS scores matching the full protocol score) (Table 2). The seated upper extremity task grouping had significantly smaller total body and arm score differences compared to the stand/walk/run task grouping (p<0.02, Friedman test) and significantly smaller leg score differences compared to the seated lower extremity task grouping (p=0.03, Friedman test). Finally, the seated upper extremity task grouping also had the lowest variance in score differences when compared to the seated lower extremity task grouping (p=0.003-0.02, F-test) and the stand/walk/run task grouping (p=0.001, F-test) across total body, arm, and leg GDRS scores. The seated upper extremity task also had a significantly lower variance of leg score differences when compared the lying down task grouping (p=0.002, F-test).

**Table 2.** Differences in GDRS scores between each task grouping and the full protocol for the body (arms+legs+trunk), arms, and legs.

|  | Task grouping GDRS score minus Full protocol GDRS score for each subject (median, range) |  |  |  |  |  |  | No. subjects with median score difference of 0 | Adjusted *p* |  |
| --- | --- | --- | --- | --- | --- | --- | --- | --- | --- | --- |
|  | 1 | 2 | 3 | 4 | 5 | 6 | 7 |  | Friedman test (differences) | F-test (variance of differences) |
| **Total Body** |  |  |  |  |  |  |  |  |  |  |
| Lying down | -1, -3-0 | 0, -5-0 | 9, 3-13 | 3, 0-5 | 0, 0-11 | 0, 0-6 | 1, -2-6 | 3 | 1.0 | 0.6 |
| Sitting – UE | 0, -1-4 | 0, -2-0 | -2, -6-1 | 0, 0-1 | 0, -7-3 | 0, 0-6 | 2, -7-7 | 5 | - | - |
| Sitting – LE | 4, -1-8 | 0, -1-7 | 0, -1-4 | 0, -2-5 | 10, 0-14 | 2, 0-9 | 7, -12-13 | 3 | 0.06 | 0.02# |
| Stand/Walk/Run | 3, -1-4 | 0, 0-6 | 10, 6-18 | 4, 2-8 | 10, -1-14 | 0, -2-0 | 10, -14-4 | 2 | 0.049* | 0.001# |
| **Arm** |  |  |  |  |  |  |  |  |  |  |
| Lying down | -1, -2-0 | 0, -1-0 | -1, -4-4 | 1, 0-2 | 0, 0-6 | 0, 0-2 | 2, 1-5 | 3 | 0.6 | 1.0 |
| Sitting – UE | 0, -1-2 | 0, -1-0 | -5, -5- -1 | 0, 0-1 | 0, -1-2 | 0, 0-0 | 0, -2-4 | 6 | - | - |
| Sitting – LE | 4, -1-4 | 0, -1-0 | -4, -7- -2 | 0, -1-0 | 5, 0-7 | 0, 0-1 | -1, -7-1 | 3 | 1.0 | 0.01# |
| Stand/Walk/Run | 3, 2-5 | 0, 0-2 | 2, -4-7 | 3, 2-6 | 4, 2-5 | 0, 0-0 | -6, -8-0 | 2 | 0.02* | 0.001# |
| **Leg** |  |  |  |  |  |  |  |  |  |  |
| Lying down | 0, -2-0 | 0, -4-0 | 6, 4-15 | 3, 0-3 | 0, -2-2 | 0, 0-4 | 0, -6-4 | 5 | 1.0 | 0.002# |
| Sitting – UE | 0, 0-2 | 0, -1-0 | 0, -1-3 | 0, 0-0 | 0, -5-3 | 0, 0-6 | 1, -5-4 | 6 | - | - |
| Sitting – LE | 0, -3-8 | 0, 0-8 | 6, 1-8 | 0, -1-4 | 5, 0-8 | 0, 0-9 | 6, -5-10 | 4 | 0.03* | 0.003# |
| Stand/Walk/Run | 0, -6-0 | 0, -3-4 | 7, 3-12 | 0, -3-4 | 6, -2-10 | 0, -2-0 | -4, -8-2 | 4 | 1.0 | 0.001# |

\* $p < 0.05$ for repeated measures Friedman test and post-hoc Dunn's test for multiple comparisons of
GDRS score differences between task groupings and the full protocol. # $p < 0.05$ for F-tests with
Bonferroni corrections for multiple comparisons between variances in GDRS score differences between
task groupings and the full protocol. Comparisons are only shown relative to the seated upper extremity
task, which was the only task that demonstrated significant differences from other task groupings. UE –
upper extremity. LE – lower extremity.

### Dystonic movement and severity features cited by experts

Across their written explanations, experts cited 12 discrete movement features they used to identify dystonia in the upper extremities, 15 for the lower extremities, 2 for the trunk, and 9 to describe dystonia severity. These codes and example expert quotes generating the codes are shown in Table 3.

**Table 3.** Codebook describing movement and severity features cited by experts when assessing dystonia across tasks.

| Movement codes by body region | Frequency cited | % of TOTAL | Example statement | Adjusted *p* |
| --- | --- | --- | --- | --- |
| **Arm (subtotal)** | **194** | **42.7%** |  | **0.04\*** |
| Wrist flexion | 38 | 8.4% | "Intermittently sustained wrist flexion" | 1.0 |
| Finger flexion | 33 | 7.3% | "occasional posturing of R fingers into flexion" | 0.7 |
| Wrist ulnar deviation | 31 | 6.8% | "ulnar deviation at wrist " | 1.0 |
| Shoulder abduction | 22 | 4.8% | "posturing of right arm into shoulder abduction" | 1.0 |
| Elbow flexion | 18 | 4.0% | "Right distal arm with flexion at elbow" | 1.0 |
| Finger abduction | 13 | 2.9% | "finger spread" | 1.0 |
| Finger extension | 13 | 2.9% | "mild left finger posturing (extension)" | 1.0 |
| Shoulder adduction | 9 | 2.0% | "Possibly right shoulder adduction" | 0.5 |
| Arm swing | 7 | 1.5% | "She is not swinging her arms with walking left greater than right." | 0.02* |
| Wrist extension | 6 | 1.3% | "When changing standing postures left wrist goes into extension" | 1.0 |
| Shoulder extension | 2 | 0.4% | "LUE posturing into shoulder extension" | 1.0 |
| Shoulder internal rotation | 2 | 0.4% | "internal rotation of right shoulder" | 1.0 |
| **Leg (subtotal)** | **243** | **53.5%** |  | **<0.0001\*** |
| Toe dorsiflexion | 38 | 8.4% | "VERY subtle - extensor posturing of the bilateral big toes, activated by voluntary arm and hand movements" | 0.03* |
| Ankle inversion | 36 | 7.9% | "Clear posturing of ankles (inversion) " | 1.0 |
| Ankle plantarflexion | 29 | 6.4% | "Bilateral dynamic ankle inversion, plantar flexion with any local or distant movement that also persists at rest. " | 1.0 |
| Hip adduction | 28 | 6.2% | "hip adduction right more than left activated by movement" | 0.8 |
| Hip flexion | 19 | 4.2% | "steps appeared dystonic with hip add / flex" | 0.002* |
| Ankle internal rotation | 16 | 3.5% | "Variably sustained inversion and internal rotation of the right ankle. " | 1.0 |
| Ankle dorsiflexion | 15 | 3.3% | "left ankle stiffening and slight movement into dorsiflexion with voluntary movement of arms" | 0.4 |
| Knee extension | 14 | 3.1% | "Possibly the knee extension on the left is dystonia. " | 1.0 |
| Hip internal rotation | 12 | 2.6% | "Variable internal rotation of hips R>L" | 0.7 |
| Knee flexion | 10 | 2.2% | "left knee flexion and hip flexion triggered by rolling" | 1.0 |
| Hip abduction | 8 | 1.8% | "He had some mild contractions of leg muscles when he moved his hands: slight hip flexion, abduction, and ankle dorsiflexion" | 0.2 |
| Hip internal rotation | 8 | 1.8% | "Internal rotation at hip (L>R)" | 1.0 |
| Ankle external rotation | 4 | 0.9% | "In both ankles, he remains at 90 degrees and is externally rotated" | 1.0 |
| Knee intorsion | 3 |  | "knee flexion/intorsion precipitated by movement" | 1.0 |
| Toe abduction | 3 |  | "fanning of the toes" | 1.0 |
| **Trunk (subtotal)** | **17** | **3.7%** |  | **-** |
| Trunk lateral deviation | 14 | 3.1% | "Has evidence of truncal involvement as well with thoracic movement to L with balancing" | - |
| Trunk extension | 3 | 0.7% | "Noted trunk hyperextension with rolling" | - |
| **TOTAL movement** | **454** | **100%** |  |  |

| codes |  |  |  |  |
| --- | --- | --- | --- | --- |
|  | Frequency cited | % of TOTAL | Example statement | Adjusted *p* |
| **Severity codes** | **324** | **100%** |  | **0.0004\*** |
| Functional impact | 67 | 20.7% | "Reaching movements with the right hand and arm were performed well so I think it does not impair function."<br>"Increase with running that appeared to contribute to impairment. " | 0.9 |
| Amplitude | 61 | 18.8% | "Dystonia was mild (small movement amplitude)" | 1.0 |
| Duration | 46 | 14.2% | "for postures with higher scores - showed sustained stiff posture"<br>"Generally lower scores as amplitude generally small and doesn't get persistently stuck in postures." | 0.1 |
| Trigger: specific task | 42 | 13.0% | "I only saw this posture during walking"<br>"I could only call the hands, which were very mild and only seen with turning" | 0.4 |
| Frequency | 29 | 9.0% | "upper limb postures were present frequently but not constantly"<br>"Ankle posturing appeared mild and intermittent" | 1.0 |
| Trigger: movement | 29 | 9.0% | "Only triggered by activity" | 0.8 |
| Rest (presence or absence of dystonia) | 26 | 8.0% | "Absence at rest - mild"<br>"Distal leg dystonia was present constantly, including during periods of relative rest" | 0.4 |
| Trigger: consistency | 16 | 4.9% | "Consistent triggering of left wrist into flexion with changing movement" | 0.1 |
| Speed | 8 | 2.5% | "There are hand postures that seem to slow movements" | 0.5 |
| **TOTAL severity codes** | **324** | **100%** |  |  |

\* $p < 0.05$ for Chi-square test with Yates correction followed by post-hoc Chi-square tests with Bonferroni
correction for multiple comparisons of code frequencies between tasks (lying down, seated upper extremity, seated lower extremity, stand/walk/run, and the full protocol). Of note, codes describing trunk movements were cited infrequently, and not at all for some task groupings, precluding further statistical analysis of this code category.

The most common codes describing dystonic movements in the upper extremities were wrist flexion (38/454 times that experts cited movement features, 8.4%), finger flexion (33/454, 7.3%), and wrist ulnar deviation (31/454, 6.8%) (Figure 2A, Table 3). The most common lower extremity dystonic movements cited were toe dorsiflexion (38/454, 8.4%), ankle inversion (36/454, 7.9%), and ankle plantarflexion (29/454, 6.4%) (Figure 2C, Table 3). The most common codes describing dystonia severity assessment were functional impact (67/324 times experts cited features that helped them gauge dystonia severity, 20.7%), movement amplitude (61/324, 18.8%), and movement duration (46/324, 14.2%) (Figure 2E, Table 3). In aggregate, characteristics of the stimuli that bring on dystonic movements (i.e. characteristics of dystonia triggers) most frequently influenced expert ratings of dystonia severity (87/324, 26.8%) (Figure 2E, Table 3). Dystonic movements that were consistently triggered by a particular stimulus or triggered across a greater number of stimuli were described as more severe than movements that were inconsistently triggered during only a single task.

**Figure 2.**
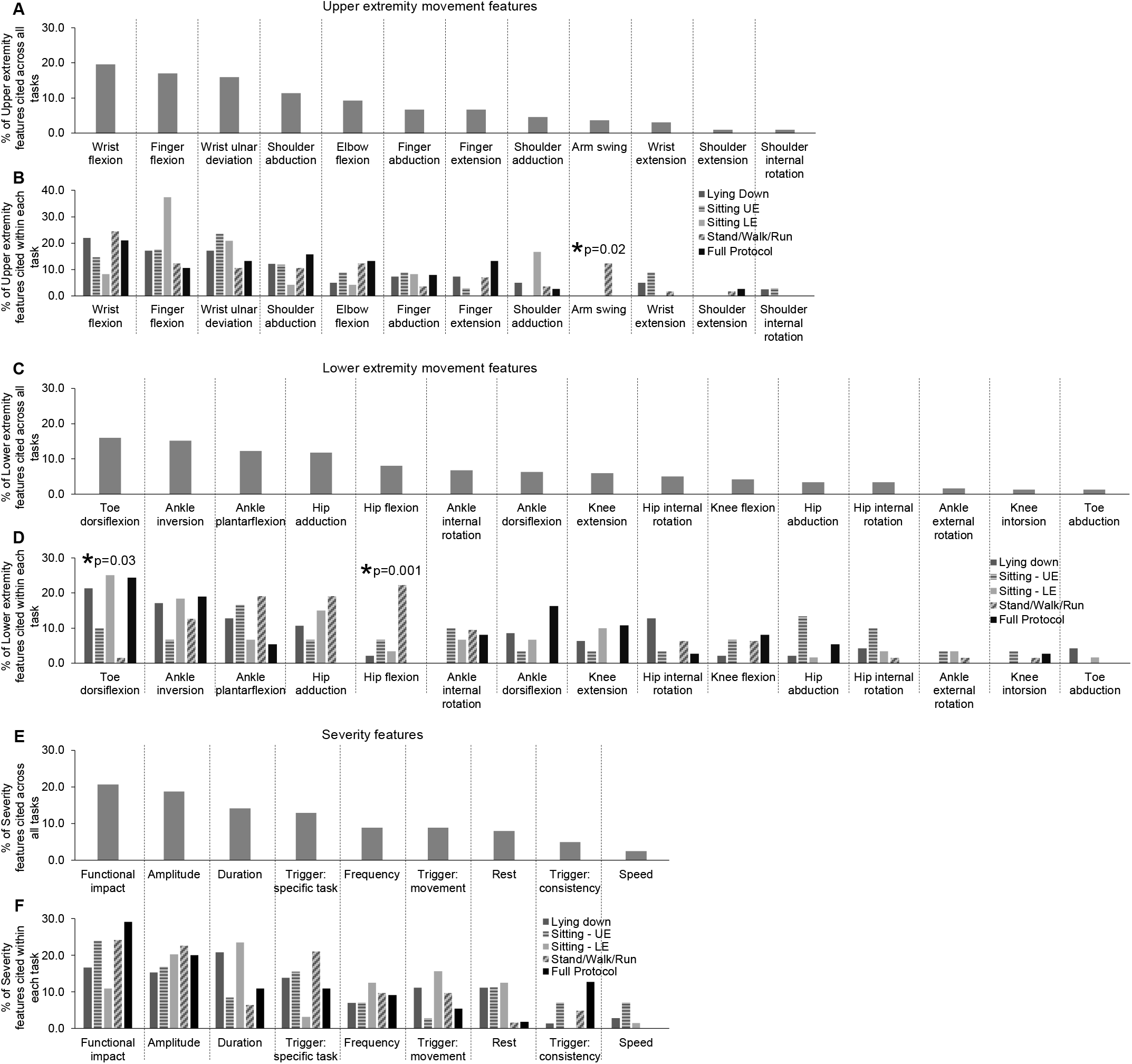
Comparison of movement and severity features cited by experts while assessing dystonia across tasks. *p<0.05 for Chi-square test with Yates correction followed by post-hoc Chi-square tests with Bonferroni correction for multiple comparisons of code frequencies between tasks. UE – upper extremity. LE – lower extremity

Codes describing trunk movements were cited infrequently, and not at all for some task groupings, precluding further statistical analysis of this code category. For the other three code categories (upper extremity, lower extremity, and severity), code frequency distributions did differ significantly between task groupings (Figure 2B, 2D, 2F, Table 2). When examining individual codes, the only significant differences between task groupings were for arm swing, toe dorsiflexion, and hip flexion (Figures 2B and 2D, Table 2). Experts disproportionately cited these codes when assessing the stand/walk/run task grouping, either significantly more often (arm swing, hip flexion) or less often (toe dorsiflexion) compared to their assessments during other task groupings.

## Discussion

Our results suggest that dystonia assessment during seated upper extremity tasks may be most comparable to dystonia assessments during a comprehensive video protocol in a small group of independently ambulatory young people with CP with mild dystonia. Our data also suggest common movement features used by experts to assess upper and lower extremity dystonia: 1) wrist and finger flexion with wrist ulnar deviation, 2) toe dorsiflexion (very likely describing a striatal toe) with plantarflexion and inversion at the ankle (i.e. equinovarus). Experts typically assessed dystonia severity using the type and consistency of the trigger and the functional impact of the movements. Enumerating these movements and severity features may provide practical guidance for how to assess dystonia clinically.^10,21^

Evaluation of a single task likely does not provide the most comprehensive assessment of dystonia. However, being vigilant for dystonia during seated upper extremity tasks may be a clinically feasible option for clinicians caring for children with CP who are looking for a screening examination or a way to grossly track dystonia severity visit to visit. Therefore, though it can be prohibitively time-consuming to use a comprehensive dystonia severity scale during every visit, grossly tracking dystonia severity during seated upper extremity tasks may be a valuable way to assess the clinical impacts of dystonia treatments over time.

It was surprising that the stand/walk/run task grouping led to greater score variability and higher dystonia severity scores when compared to the full protocol, particularly because we have previously demonstrated that leg adduction amplitude and variability linearly correlate with expert assessments of leg dystonia severity during gait.^22^ It may be more difficult to distinguish between dystonia and other sources of motor disability in individuals as they walk (e.g. spasticity, weakness, ataxia, etc.), leading to variable attribution of abnormal posturing observed during walking to dystonia.^23,24^ Differences between these motor symptoms may be clearer in tasks with more periods of relative rest to distinguish between static versus triggered symptoms.^25,26^ Indeed, there were fewer mentions of “rest” as a feature used to assess dystonia severity when assessing the stand/walk/run task grouping compared to other task groupings, though this difference did not reach significance. In addition, noting that “functional impact” was a common expert-cited feature used to assess dystonia severity, it is possible that functional impact is rated higher when assessing gait alone. The stand/walk/run task grouping is the only grouping that exclusively assessed tasks individuals may otherwise routinely do. For example, though a seated hand open-close task can be valuable for clinically assessing dystonia severity, it is not a task that is regularly performed as a part of daily life. Therefore, the functional impact of dystonia may be rated by experts more highly when assessing only a highly functionally relevant task like gait.

### Limitations

Limitations of this study primarily center around population demographics and sample size. To assess stand/walk/run as a task grouping, we had to limit participant eligibility to those who were independently ambulatory. This precludes generalizability of these results to those who use primarily use wheelchairs for mobility. Because dystonia is more common and likely more severe in people with greater gross motor functional limitations, recruiting independently ambulatory individuals also limited our ability to recruit people with moderate to severe dystonia. To mirror the desired real-life application of video protocols for dystonia assessment, we attempted subject recruitment and filming during routine clinical care. Many families declined participation noting that recording the full video protocol took up to an hour of time. Though this dramatically limited our sample size, it does highlight the importance of creating a feasible video protocol for dystonia assessment during routine clinical care both for efficient use of clinician time and family time. Also of note, participants had to have the cognitive and physical stamina to record a video protocol for one hour, which again likely limited us to recruiting participants with relatively mild dystonia and high motor function. In sum, this study must be replicated in a larger and more functionally diverse group of people with CP.

Additional limitations include that our videos were only generated from subjects seen at a single center, though this was a large tertiary-care referral center and though our dystonia assessment experts come from five different centers across three countries. Also of note, to ensure the anonymity of our participants who provided this video as a part of routine clinical care, we had to blur their faces. This precluded assessment of face and neck dystonia which should be addressed in future work. Finally, it will be valuable to assess the generalizability of these results to adults and people with generalized dystonia from different etiologies.

### Conclusions

In ambulatory young people with CP, this preliminary work suggests that assessing for dystonia during seated upper extremity tasks (compared to seated lower extremity tasks or standing/walking/running) may provide a useful and feasible approximation of dystonia severity during routine clinical care. Additionally, this work demonstrates key movement features experts use to identify dystonia and assess its severity in ambulatory people with CP – findings that can provide immediate clinical utility to guide dystonia assessment. Though this work is preliminary, it highlights the incredible difficulty in having people with CP complete the exhaustive list of tasks currently required to assess dystonia using existing dystonia rating scales. We must move towards clinically feasible dystonia assessment to facilitate clinical care and this work marks an important step towards that necessary goal. Future work can assess the generalizability of these findings to a larger population of people including those with moderate-severe dystonia, those who primarily use wheelchairs for mobility, subjects from other centers, adults, and people with generalized dystonia due to other etiologies.

## Acknowledgement

The authors acknowledge the participants and their families who, during routine clinical care, sacrificed an additional hour of time to record comprehensive video protocols and selflessly demonstrated the incredible burden on families in using these comprehensive protocols to assess dystonia severity.

## Author contributions

Alyssa Rust: Data curation, Formal analysis, Investigation, Writing – review and editing

Leon Dure: Investigation, Writing – review and editing

Darcy Fehlings: Investigation, Writing – review and editing

Toni Pearson: Investigation, Writing – review and editing

Roser Pons: Investigation, Writing – review and editing

Jonathan W. Mink: Investigation, Writing – review and editing

Bhooma R. Aravamuthan: Conceptualization, Data curation, Formal analysis, Funding acquisition,, Investigation, Methodology, Project administration, Resources, Software, Supervision, Validation, Visualization, Writing – original draft, Writing – review and editing

## Ethical compliance statement

The Washington University Institutional Review Board approved this project (Approval Number: 202306016; Date of Approval 06/05/2023). Subjects provided verbal and written consent for videos to be recorded during routine clinical care. We confirm that we have read the Journal’s position on issues involved in ethical publication and affirm that this work is consistent with those guidelines.

## Supplemental Methods

Comprehensive video protocol.

